# Machine Learning and Bioinformatics Unravel Gene Signatures of Coronary Artery Disease Comorbidity with Periodontitis

**DOI:** 10.1101/2024.09.18.24313934

**Authors:** Nannan Yang, Wenqian Yang, Kun Wang, Liguo Tan, Hao Zhang, Zhen Huang, Qi Chen, Huanhuan Xing, Ying Jin, Wenting Liu, Shaobing Wang

## Abstract

**Background:** Emerging evidence suggests a complex interplay between periodontal disease (PD) and coronary artery disease (CAD) risk. The novel insights into the shared pathogenesis of PD and CAD will potentially inform future therapeutic strategies. This study aimed to identify signature genes implicated in the progression of PD to CAD.

**Methods:** Gene expression data from NCBI GEO datasets, GSE10334 and GSE66360, associated with both PD and CAD datasets were analyzed to pinpoint differentially expressed genes (DEGs), followed by weighted gene co-expression network analysis (WGCNA) to identify key modules. Functional enrichment analysis of common DEGs was conducted. Four machine learning algorithms were employed to construct predictive models, and the optimal model was selected for subsequent feature genes selection. Using GSE6751 and GSE71226 as validation cohort, receiver operating characteristic (ROC) curves and nomograms were generated for diagnostic performance assessment and risk prediction. Furthermore, immune cell infiltration patterns were assessed using the CIBERSORT (Cell-type Identification By Estimating Relative Subsets Of RNA Transcripts) algorithm. Finally, RNA-sequencing (RNA-seq) of 5 clinical samples vs. 5 controls was performed to validate the identified genes and explore their potential as biomarkers for early diagnosis and prevention of comorbid periodontitis and CAD.

**Results:** Analysis of the GSE10334 and GSE66360 datasets revealed 48 common Differentially Expressed Genes (DEGs) associated with CAD and PD. Gene Ontology (GO) and Kyoto Encyclopedia of Genes and Genomes (KEGG) pathway enrichment analyses of these DEGs highlighted significant overrepresentation of pathways related to inflammatory responses and immune cell trafficking, including response to lipopolysaccharides, molecules of bacterial origin, neutrophil migration, bone marrow leukocyte migration, and CXCR chemokine receptor binding. Additionally, pathways involved in lipid metabolism and atherosclerosis, such as the NF-κB signaling pathway, IL-17 signaling pathway, and TNF signaling pathway, were also enriched. Five genes (*FOS, MME, PECAM1, RGS1,* and *VNN2*) emerged as potential signature genes, demonstrating strong predictive ability with an area under the curve (AUC) greater than 0.7 on the machine learning algorithms. CIBERSORT analysis suggested a potential role of these signature genes in modulating immune cell infiltration. To further validate these findings, RNA-seq on clinical samples confirmed significant upregulation of *FOS*, *VNN2*, *PECAM1*, and *MME* genes in patients with both CAD and PD.

**Conclusion:** This study identified five signature genes that were significantly associated with immune cell dysregulation, where four of them were verified on clinical samples. These genes hold promise for the development of a nomogram-based approach for early diagnosis of both periodontitis and coronary artery disease, potentially informing future research directions for improved diagnosis and treatment strategies in these prevalent conditions. Notably, the prominent upregulation of *FOS* suggests its potential as a key target for future investigations. These insights hold significant implications for improving prevention and diagnostic strategies for individuals affected by both PD and CAD.

## 1 Introduction

Periodontal disease is commonly characterized as an inflammatory condition affecting the tissues surrounding and supporting the teeth, including the gums and periodontal tissues. It is a complex chronic inflammatory disease with a prevalence ranging from 20% to 50% of the global population in severe cases, according to the World Health Organization. Over the past 30 years, the incidence, prevalence, and disability rates of periodontal disease have been on the rise. This prevalent disease burden has seen PD rise to rank among the world’s 15 most common conditions, with a concerning association with increased adult mortality rates [1]. Formal genetic studies have confirmed a significant hereditary component, attributing roughly half of the population variation in chronic PD to genetic factors [2]. Similarly, cardiovascular diseases (CVDs) represent a leading cause of global mortality. coronary artery disease, the most prevalent form of CVD, is characterized by narrowed or blocked coronary arteries due to atherosclerosis or spasm, leading to reduced oxygen supply (ischemia) and potential tissue death (infarction) within the heart muscle [3]. Globally, CAD claims over 7 million lives annually. In China alone, 2020 data revealed concerning mortality rates: 126.91/100,000 in urban and 135.88/100,000 in rural areas, with a continued upward trend [4].

Growing research interest in recent decades has focused on the link between periodontal disease and cardiovascular disease. Numerous studies have established a significant association between periodontitis and coronary artery disease, evident at the bacterial, genetic, and various other risk factor levels [5]. This association is supported by two key observations: first, both diseases share common susceptibility factors, and second, PD acts as a risk factor, potentially initiating atherogenesis, promoting plaque maturation, and contributing to its instability [5]. Furthermore, periodontitis shares pathogenic pathways with CVD. Periodontal bacteria trigger the release of pro-inflammatory mediators, both locally and systemically, which directly worsen CAD and contribute to coronary plaque rupture [6]. Kyari et al. (2014) employed data from the 2013-2014 National Health and Nutrition Examination Survey (NHANES) in a cross-sectional study involving 2,830 adults aged 30 or older. Their findings indicated that individuals with fair/poor gingival health had a 2.17-fold increased risk of developing CVD compared to those with good/very good gingival health (95% CI, 0.98-4.79, *P* = 0.055) [7]. Additionally, a large cohort study demonstrated that both new-onset and existing periodontitis are linked to an increased risk of coronary artery disease. This study also identified a graded relationship between tooth loss and poorer patient outcomes, including stroke, cardiovascular events, and overall mortality, specifically in individuals with stable coronary artery disease [8]. Joshi et al. (2021) reviewed data from 14 studies, demonstrating the presence of periodontal bacterial DNA in atherosclerotic plaque specimens from patients with coronary artery disease. Notably, porphyromonas gingivalis DNA exhibited significantly higher expression in these plaques compared to healthy tissue (mean prevalence 0.4; 95% CI, 0.24-0.56; *P* < 0.001) [9]. Further evidence comes from Stryjewska et al. (2019) who studied 128 individuals, including 68 patients with acute myocardial infarction (AMI) and 60 healthy controls. They found a significantly higher rate of fungal colonization in the AMI group (50% vs. 25%), with Candida albicans being the predominant fungus (44% vs. 17%). Importantly, oral fungal colonization (OR 3.0; 95% CI 1.4-6.4), particularly by C. albicans (OR 3.7; 95% CI 1.9-9.1), emerged as a strong predictor of AMI. Collectively, these findings suggest that periodontal disease and dental caries may act as reservoirs for pathogenic microorganisms. These microbes can potentially migrate through the bloodstream and disseminate to various organs, including atherosclerotic plaques, potentially increasing plaque instability and rupture [10].

This study employed weighted gene co-expression network analysis to investigate the correlation between modular signature genes (highly interconnected sets of genes) and the occurrence of periodontitis and coronary artery disease. Differentially expressed genes associated with both diseases were identified using GEO2R. Genes showing overlap between WGCNA modules and DEGs were selected for further analysis. Subsequently, the potential link between periodontitis and coronary artery disease was explored through signature gene identification and immune infiltration analysis, and RNA-sequencing (RNA-seq) was performed to validate the identified genes and explore their potential as biomarkers for early diagnosis and prevention of comorbid periodontitis and CAD.to provide a new reference for the diagnosis and treatment of patients with coronary artery disease secondary to periodontitis.

## 2 Methods

### 2.1 Raw data

Gene expression data were obtained from the Gene Expression Omnibus (GEO) database (https://www.ncbi.nlm.nih.gov/geo/) using the accession numbers GSE10334 [12] and GSE66360 [13]. The GSE10334 dataset includes 247 samples derived from 90 patients. These samples encompass 183 tissues affected by disease and 64 healthy control tissues from the same patients. The GSE66360 dataset consists of isolated circulating endothelial cells from 49 patients diagnosed with acute myocardial infarction and 50 healthy individuals.

### 2.2 WGCNA network construction and module identification

The WGCNA package in R [14] was employed to construct co-expression networks and identify co-expressed gene modules. This process involved several steps. First, samples underwent hierarchical clustering to identify outliers, which were subsequently removed alongside any unqualified genes using the goodSamplesGenes function. Second, the soft-thresholding power (β) was calculated using the R function pickSoftThreshold and converted into a topological overlap matrix (TOM). Third, based on the TOM, the network’s interconnectivity was established, and the dynamic tree-cutting algorithm was applied to identify co-expressed gene modules. Fourth, modules are associated with clinical features by calculating gene saliency and module affiliation. The genes involved in the corresponding modules were used for subsequent analysis. Finally, the feature gene network was visualized.

### 2.3 Identification of common DEGs

GEO2R (www.ncbi.nlm.nih.gov/geo/ge2r) is an online deg analysis tool based on the Limma software package [15]. An adjusted P-value < 0.01 and |logFC| ≥ 1 were defined as thresholds for screening DEGs. We extracted the genes most associated with periodontitis and coronary artery disease from the WGCNA module and cross-analyzed the two gene lists for common DEGs.

### 2.4 Enrichment analyses of common DEGs

To further understand which biological functions these common DEGs are involved in, we performed gene enrichment analysis using the KOBAS 3.0 database, including gene ontology and Kyoto Encyclopedia of Genes and Genomes pathways [16]. Adjusted P-values < 0.05 was considered significant.

### 2.5 Machine learning algorithms for identifying feature genes

Machine learning models are constructed using the filtered DEGs. We used four machine learning methods to construct the model respectively:

1. Random Forest (RF): This versatile ensemble method combines multiple decision trees, each built independently from random subsets of the data [17, 18]. Individual trees learn and predict independently, with the final prediction being the average of all tree predictions.
2. Support Vector Machine Recursive Feature Elimination (SVM-RFE): This discriminative classifier iteratively removes the least informative features to identify the most relevant ones for classification [19]. The model is trained using labeled samples, and test samples are classified based on the optimal separation hyperplane.
3. XGBoost: This powerful ensemble method leverages gradient boosting to create a robust model by combining multiple weak decision trees [20]. XGBoost facilitates a meticulous comparison of classification errors and model complexity.
4. Generalized Linear Model (GLM): This flexible method extends the linear model by incorporating a link function to model the relationship between the response variable and the linear combination of predictor variables [21]. We compared the performance of these four methods to identify the one with the highest accuracy for screening disease signature genes. The “DALEX” package was used to interpret the models, allowing us to visualize the residual distributions and feature importance for each method. Performance was further evaluated by visualizing the receiver operating characteristic curves using the “qROC” package. A larger area under the curve indicates higher model accuracy.

### 2.6 Receiver operating characteristics curve and nomogram construction

To assess the diagnostic potential of identified characteristic genes, their expression levels in coronary artery disease and periodontitis groups were compared. The area under the receiver operating characteristic curve was calculated for each gene, along with its 95% confidence interval, to estimate diagnostic accuracy. To mitigate bias, datasets GSE6751 and GSE71226 were employed as validation cohorts. Only genes exhibiting an AUC > 0.7 in both the original dataset and both validation sets were included for further analysis. Subsequently, nomograms were constructed using the RMS R package [22] to facilitate clinical decision-making.

### 2.7 Immune Infiltration Analyses

CIBERSORT is a bioinformatics tool that estimates the relative proportions of various cell types within complex tissue samples [23]. We utilized CIBERSORT to evaluate immune cell infiltration in the coronary artery disease and periodontitis groups. This algorithm offers a comprehensive assessment of immune cell abundance by profiling 24 distinct immune cell types. The proportion of each immune cell type in each sample was visualized using histograms. Additionally, heatmaps depicting the correlations between different immune cell types within the CAD and PD groups were constructed using the “corrplot” R package [24]. We further employed t-test to compare immune cell infiltration levels between disease and control groups. Finally, Spearman correlation analysis was performed to explore potential associations between immune cell populations and signature genes.

### 2.8 Clinical validation of RNA-seq analysis

Blood samples and clinical data of 5 patients with both PD and CAD, and another 5 patients with only CAD, were collected at Shiyan People’s Hospital from May 2024 to July 2024. In order to obtain a homogeneous sample with patients enrolled in a specific age group (35-75 years) and equal representation of men and women, stratified random sampling was performed by dividing patients who met the study criteria into three age strata of 35-50, 51-65, and 66-75 years to reduce possible bias related to gender or different ages. The patients included in the study were categorized into CAD group, PD+CAD group.

The exclusion criteria were (1) those who had taken antibiotics or hormonal drugs within six months; (2) women who were in pregnancy or breastfeeding; (3) systemic diseases and history of drug allergy; (4) those who had a history of treatment for periodontal diseases within one year; (5) those who suffered from renal and thyroid diseases; and (6) those who had used non-steroidal anti-inflammatory drugs within three months. The study experimental design and workflow were approved by the Science and Technology Ethics Review Committee of Hubei Medical College (reference number: 2024-RE-26). Written consent was obtained from patients before inclusion in the study.

All patients included in the study had 5 ml of venous blood collected on admission, placed in an anticoagulation tube containing 3.2% sodium citrate, left to stand for 5 min at room temperature, centrifuged at 3500 r/min, the precipitate extracted, 1 ml of Trizol was added and mixed, and the peripheral blood samples was immediately frozen and stored on dry ice. RNA extraction of the samples was performed using Tianmo#TR205-200 kit. Extracted total RNA was tested for RNA integrity using an Agilent Bioanalyzer 2100 (Agilent technologies, Santa Clara, CA, US) test with a Qubit® 3.0 Fluorometer (Life Technologies, CA, USA) and a Qubit® 3.0 Fluorometer (Life Technologies, CA, USA) and Nanodrop One spectrophotometer (Thermo Fisher Scientific Inc, USA) were used to determine the concentration and purity of total RNA.

Total RNA was extracted for microarray analysis from venous blood of 10 patients who met the study inclusion criteria. The microarray study was performed using Affymetrix GeneChip®Human Genome U133 Plus 2.0 Array for standard operating procedures and quality control of mRNA. Samples from different groups were randomized to prevent batch effects and sample processing was performed simultaneously to minimize variation. Quality control of microarray data was performed using the Transcriptome Analysis Console (TAC). For each mRNA expression probe set, it was then normalized using the TMM (trimmed mean of M values) algorithm (https://www.ncbi.nlm.nih.gov/COG/). The median polish after quantile normalization was used to remove probe affinity effects when calculating probe set summaries. Expression values were log2 of signal intensity. low expression probe sets were removed if they were not expressed in more than 25% of the samples. The expression level of each gene was estimated by calculating the median expression level of all probes within the coding region of that gene. Residual normalized gene expression was fitted to a regression model using the R package “limma”. A mitigated t-test was used to determine the deg between the CAD and PD+CAD patient groups. Genes with P-value < 0.05 and at least a 1-fold change (FC) were determined to be significant DEGs.

Total RNA sequencing libraries were prepared using an Illumina TruSeq Stranded Total RNA Library Prep kit to deplet cytoplasmic rRNA according to the manufacturer’s instructions. All cDNA libraries were QC using the Agilent DNA1000 Kit (Agilent) prior to next-generation sequencing. samples were sequenced on an Illumina NovaSeq 6000 to a depth of approximately 50 million pairs of end-reads per biological sample. FastQ was utilized for quality control of raw reads. Pairwise end-reads were aligned to the human reference genome GRCh38 using the STAR comparator with default parameters. Gene counts were calculated using featureCounts [25]. Gene counts were normalized for “limma” default parameters. And differential gene expression (DE) analysis was performed using “limma” package. The significant deg called with P-value < 0.05 and |logFC| >1. Graphs were generated using R (version 4.3.1).

## 3 Results

### 3.1 WGCNA network construction and module identification

WGCNA was employed to identify co-expressed gene modules potentially associated with disease. Before module identification, samples were clustered to detect and remove outliers. No significant outliers were identified in the GSE10334 (Fig. 2A) or GSE66360 (Fig. 2C) datasets. Within the WGCNA framework, an optimal soft-threshold power (β) of 11 was determined for the GSE10334 dataset (Fig. 2B), while a β of 8 was optimal for the GSE66360 dataset (Fig. 2D). This analysis resulted in the identification of nine modules in the GSE10334 dataset and seven modules in the GSE66360 dataset. Correlation analysis revealed a strong negative correlation between the brown module and periodontitis (r = −0.55) (Fig. 2E). Similarly, the grey module exhibited a strong negative correlation with coronary artery disease (r = −0.65), while the brown was positively correlated with coronary artery disease (r = 0.61) (Fig. 2G). Moreover, the correlation of module membership in brown (r = 0.83) and gene significance for PD samples was observed, as well as grey module membership (r = 0.66) (Figure. 2F, H).

**Figure 1.**
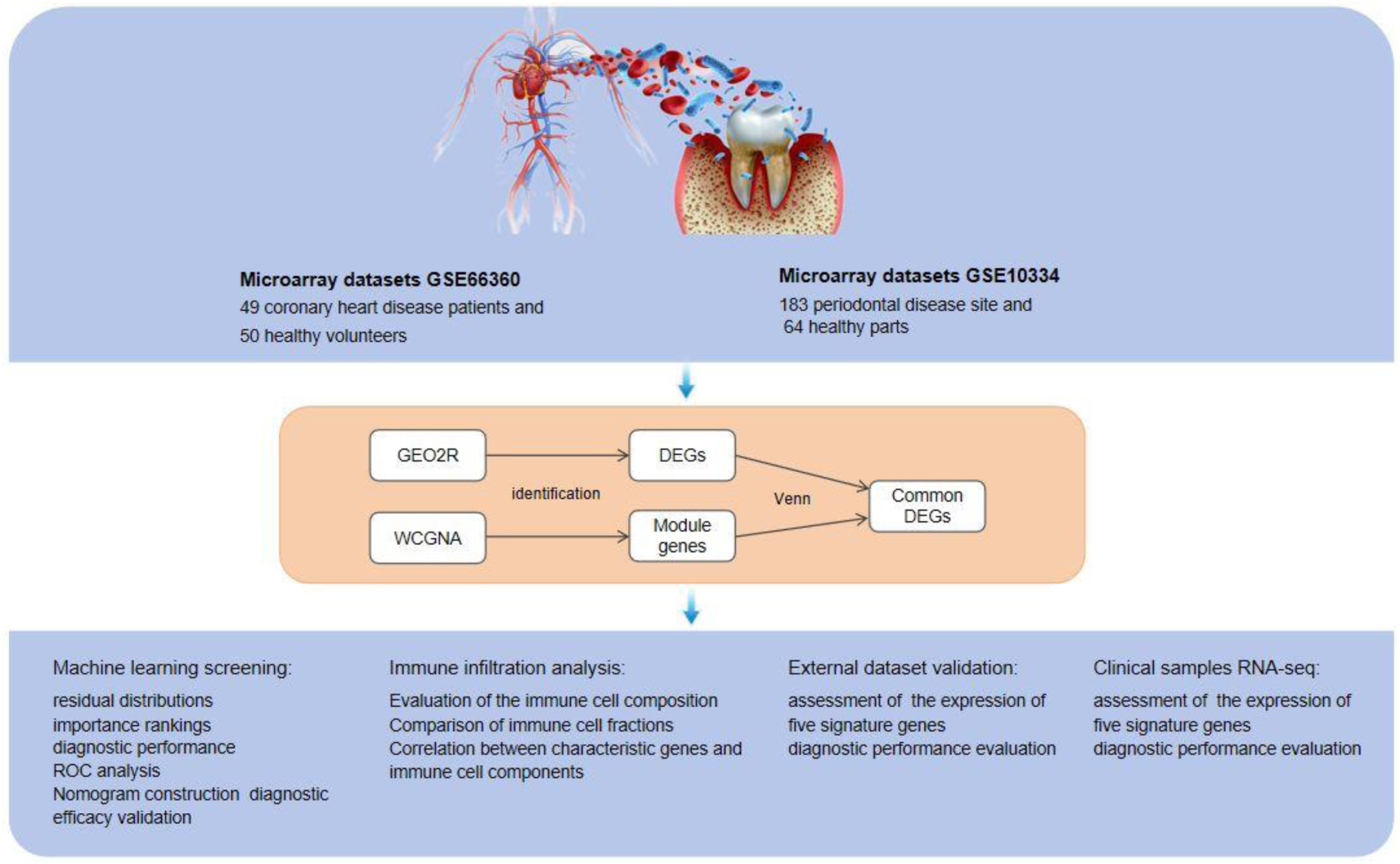
Research design flow chart. In this study, we used WCGNA to explore the correlation between modular signature genes and the occurrence of periodontitis and coronary artery disease, while DEGs for periodontitis and coronary artery disease were obtained using GEO2R. The overlapping genes between modular genes and DEGs were used for further analyses. Subsequently, the optimal machine learning model was selected to identify the signature genes of the two diseases and validate the accuracy of the model, and the expression of the signature genes was verified in the validation dataset. Immune infiltration analysis and gene-immune cell correlation analysis were then performed to demonstrate that they were significantly associated with immune cell dysregulation. Finally, RNA-sequencing of clinical samples was performed to validate the identified genes and explore their potential as biomarkers for early diagnosis and prevention of co-morbid periodontitis and CAD.

**Figure 2.**
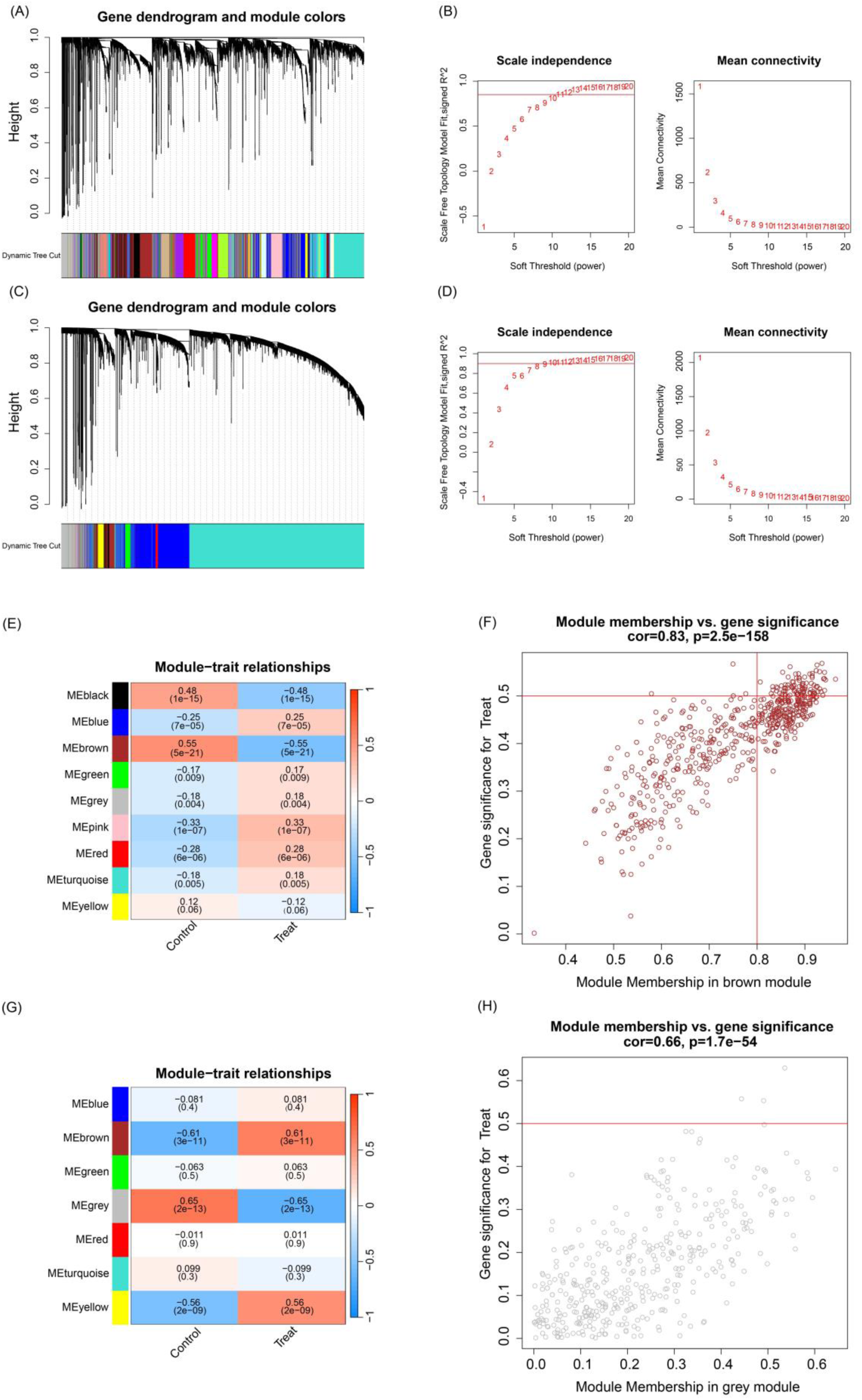
(A, C) Clustering dendrogram of samples based on their Euclidean distance in GSE10334 and GSE66360. (B, D) Determination of soft-thresholding power for GSE10334 and GSE66360. (E, G) Heatmap of the correlation between module genes and the occurrence of periodontitis and coronary artery disease. Modules with different colors represent different gene modules, and the numbers in the modules represent the correlation between the module and the phenotype. (F, H) The scatter plots of module membership and gene significance for PD and CAD respectively.

### 3.2 Identification of common DEGs

Differential expression analysis using GEO2R identified 517 and 687 DEGs in the GSE10334 and GSE66360 datasets, respectively (Figure. 3A, B). We then focused on genes within WGCNA modules that exhibited strong associations with periodontitis and coronary artery disease. Genes from these disease-associated modules were intersected with the GEO2R-identified DEGs to identify commonly dysregulated genes in both diseases. Utilizing Venn diagram analysis, this approach revealed 48 DEGs shared between periodontitis and CAD (Figure. 3C).

**Figure 3.**
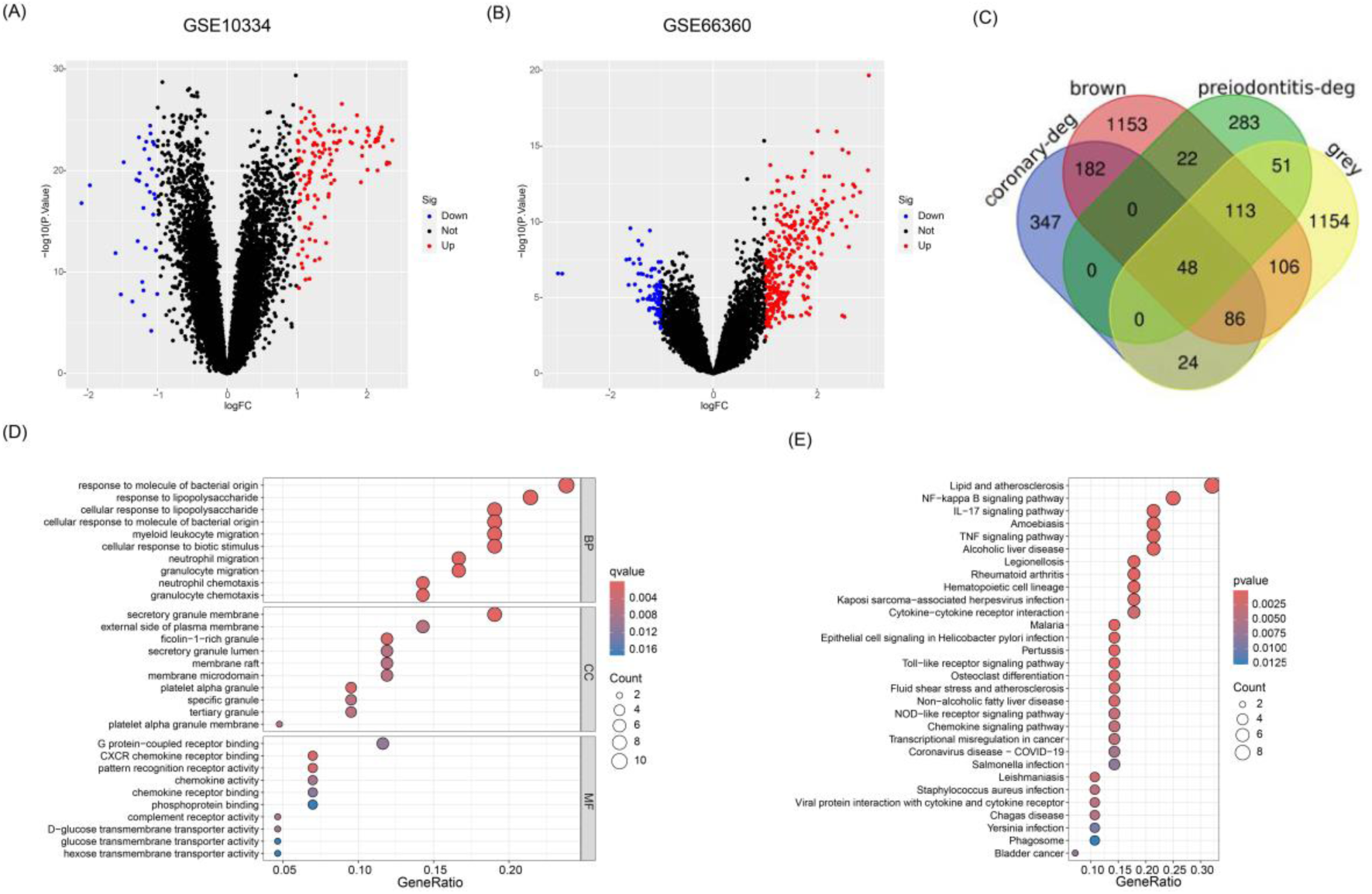
(A) The volcano map of GSE10334. (B) The volcano map of GSE66360. Red dots represent up-regulated genes, blue dots represent down-regulated genes and grey dots represent genes with no significant difference. (C) Venn diagram shows that 48 common DEGs in GSE10334 and GSE66360. (D) Enrichment result of common DEGs GO term; (E) enrichment result of common DEGs KEGG pathway. Adjusted P-value < 0.05 was considered significant. The ordinate represents the enriched term, and the abscissa represents the proportion of genes involved in the term. The size of the dots represents the number of genes, and the color of the dots represents the P-value.

### 3.3 Analysis of the functional characteristics

Gene Ontology enrichment analysis revealed that commonly dysregulated genes were significantly enriched in biological processes including response to lipopolysaccharide, response to bacterial-derived molecules, bone marrow leukocyte migration, and neutrophil migration (Fig. 3D). Cellular component enrichment highlighted secretory granule membranes, secretory granule lumen, and plasma membrane. Furthermore, enriched molecular functions included CXCR chemokine receptor binding, pattern recognition receptor activity, and phosphoprotein binding. KEGG pathway analysis identified significant enrichment of DEGs in pathways associated with lipids and atherosclerosis, the NF-κB signaling pathway with IL-17, and the TNF signaling pathway (Fig. 3E).

### 3.4 Candidate signature genes of CAD and PD using machine learning

We utilized the identified intersection genes to train four machine-learning models. The “DALEX” package in R was employed to visualize the residual distributions of each model within the training set. Subsequently, feature importance analysis was performed to rank the top ten genes in each model based on their Root Mean Squared Error (RMSE). Finally, the diagnostic performance of all four models in the training set was evaluated using ROC curves generated through 5-fold cross-validation.

Analysis of the residual distributions (Fig.4A, B) revealed that the Support Vector Machine and XGBoost models exhibited lower sample residuals, indicating better model fitting. Furthermore, the XGB model demonstrated the largest area under the ROC curve (AUC) in Fig. 4D, signifying superior prediction accuracy. Consequently, the top five most important genes (*FOS, VNN2, PECAM1, MME, RGS1*) identified by the XGB model were selected as predictor variables for further analysis (Fig. 4C). To assess generalizability, the XGB model was validated on independent datasets (GSE106090, GSE179789) from the validation set. The results depicted in Fig. 4E and Fig. 4F confirm that the XGB model retains good predictive ability in these external datasets.

**Figure 4.**
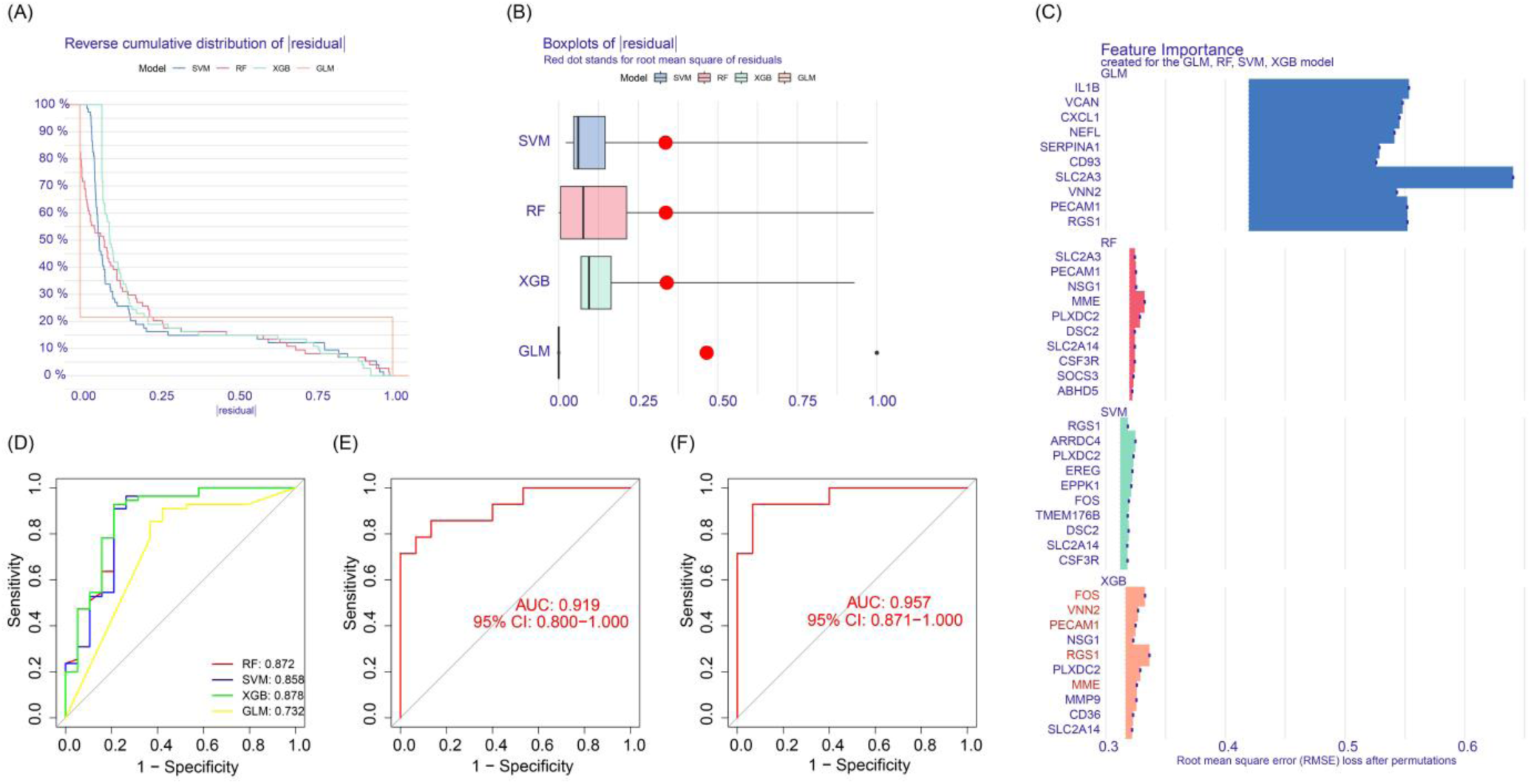
Construction and evaluation of SVM, RF, XGB, and GLM machine learning models. (A) Cumulative residual distributions of samples computed by SVM, RF, XGB, and GLM machine learning models respectively. (B) Box line plots of sample residuals based on SVM, RF, XGB, and GLM machine learning models, with the red dots representing the root-mean-square (RMS) of the residuals. (C) The importance scores of the top ten genes obtained by SVM, RF, XGB, and GLM. (D) ROC analyses of the four machine learning models based on the 5-fold cross-validation in the training set. (E, F) ROC analysis of the XGB model in the 2 validation sets.

### 3.5 Validation of biomarker diagnostic value and nomogram construction

In the external validation set (GSE106090), all five identified signature genes exhibited significantly higher expression levels in periodontitis patients compared to healthy controls (Fig. 5A-E), suggesting their potential involvement in periodontitis pathogenesis. The AUC values of the receiver operating characteristic (ROC) curves for these genes ranged from 0.704 (*FOS*) to 0.891 (*PECAM1*) (Fig. 5F-J), all exceeding 0.7, which indicates good diagnostic potential.

**Figure 5.**
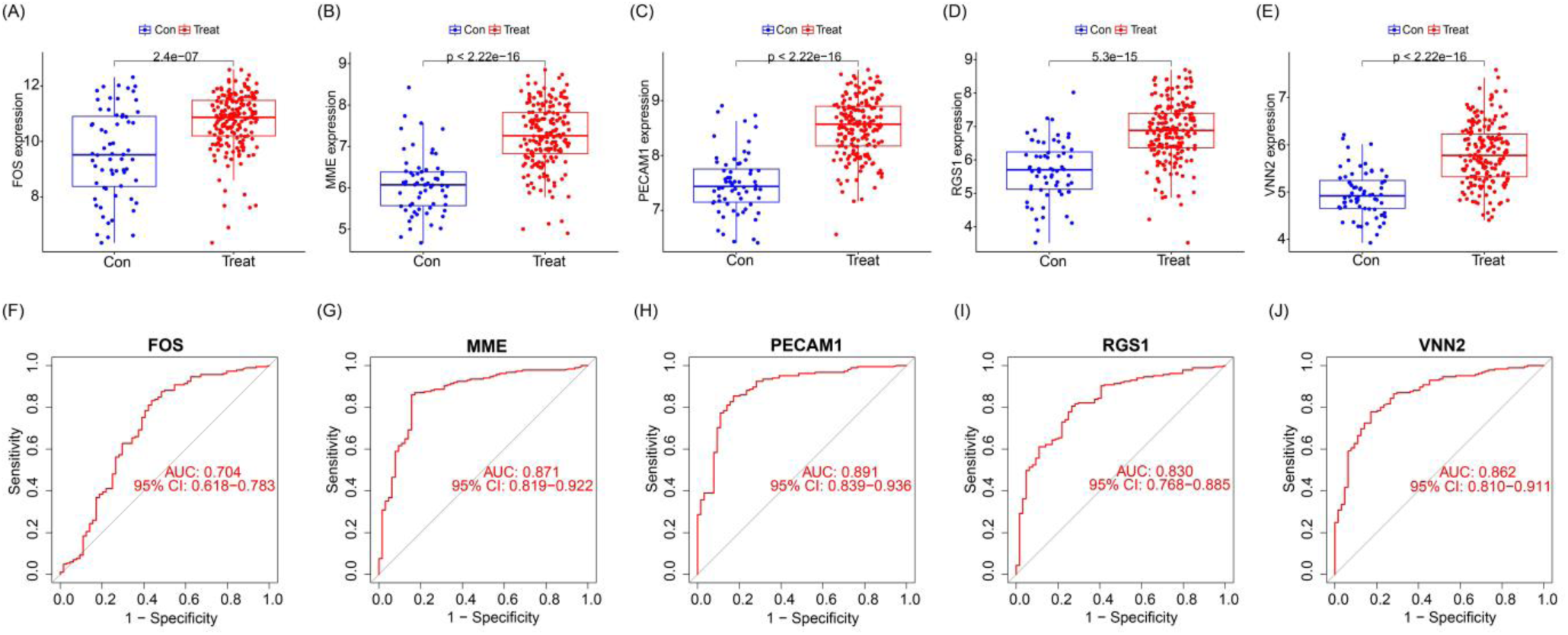
Expression of feature genes in GSE106090. (A-E) Expression of signature genes in periodontitis patients and healthy population. (F-J) ROC showing diagnostic performance of signature genes.

We further evaluated the diagnostic efficacy of these genes for coronary artery disease prediction in the validation set (GSE179789). Consistent with the periodontitis findings, all genes displayed higher expression in CAD patients compared to controls (Fig. 6A-E). The AUC values of the ROC curves for these genes ranged from 0.710 (*PECAM1*) to 0.861 (*RGS1*) (Fig. 6F-J), again exceeding 0.7, suggesting good diagnostic performance for CAD as well. Collectively, these results indicate that the identified signature genes hold promise as diagnostic biomarkers for both periodontitis and coronary artery disease.

**Figure 6.**
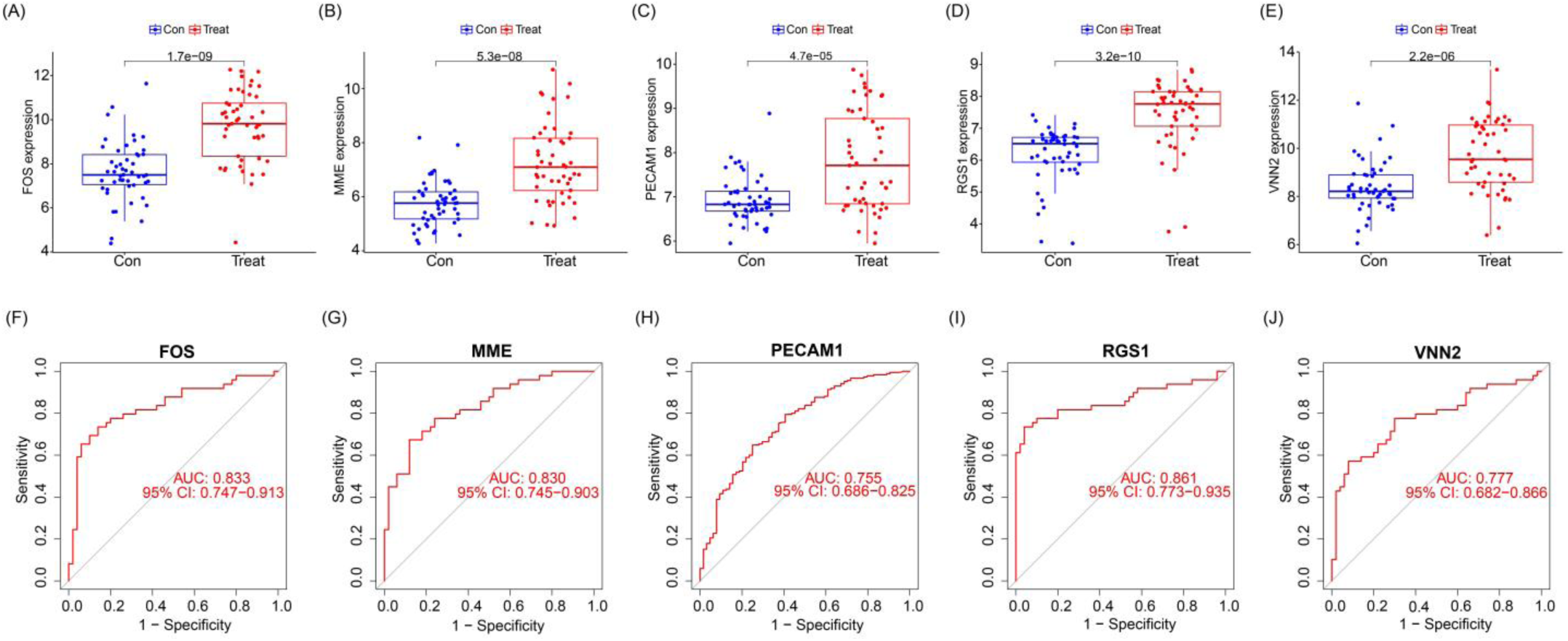
Performance of signature genes in GSE179789. (A-E) Expression of signature genes in patients with coronary artery disease and healthy population. (F-J) ROC showing diagnostic performance of signature genes.

To gain insights into the XGB model’s predictions, we employed a nomogram (Fig. 7A). This graph depicts the estimated risk of developing coronary artery disease compared to periodontitis for a given individual. The expression level of each signature gene translates to a corresponding score, and the sum of these five scores generates a composite score. This composite score serves as an indicator of disease prevalence for a specific sample. The calibration curve (Fig. 7B) demonstrates good agreement between the actual risk of developing the disease and the model’s predicted risk, highlighting minimal error. Additionally, decision curves (Fig. 7C) support the high accuracy of the nomogram for risk prediction.

**Figure 7.**
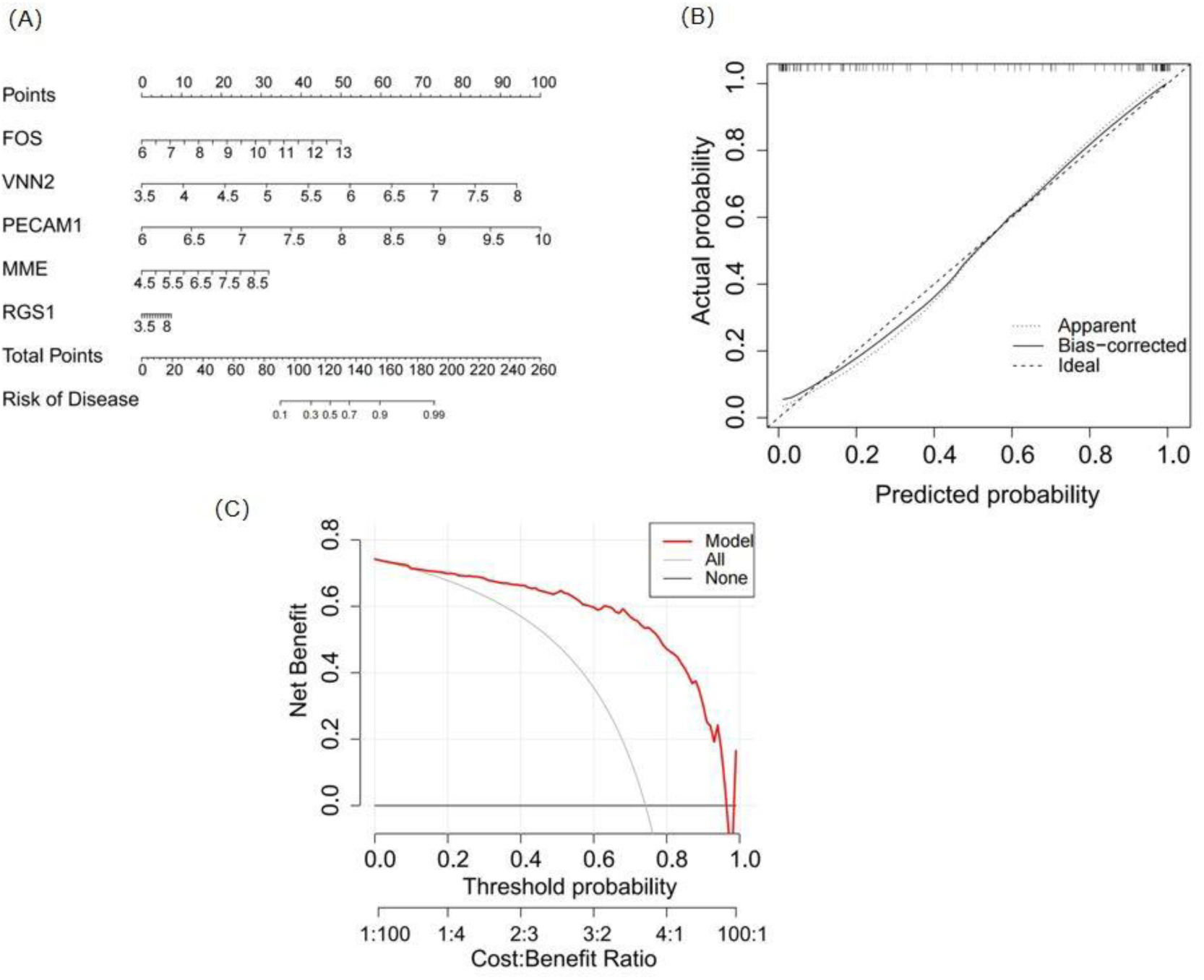
Nomogram construction and diagnostic efficacy validation. (A) A nomogram was constructed based on five selected feature genes, each corresponding to a score, and the total score of the five feature genes was used to predict the risk of periodontitis and coronary artery disease. (B-C) Calibration curves (B) and decision curves (C) were used to assess the predictive efficiency of the XGB model.

### 3.6 Immune cell infiltration analysis

Evaluation of the immune cell composition in periodontitis and coronary artery disease showed significant differences in immune cell profiles between the diseased and control groups (Fig. 8A, C). In patients with periodontitis and coronary artery disease, eosinophils, activated memory CD4+ T cells, and resting dendritic cells were highly positively correlated with B cells (Fig. 8B, D).

**Figure 8.**
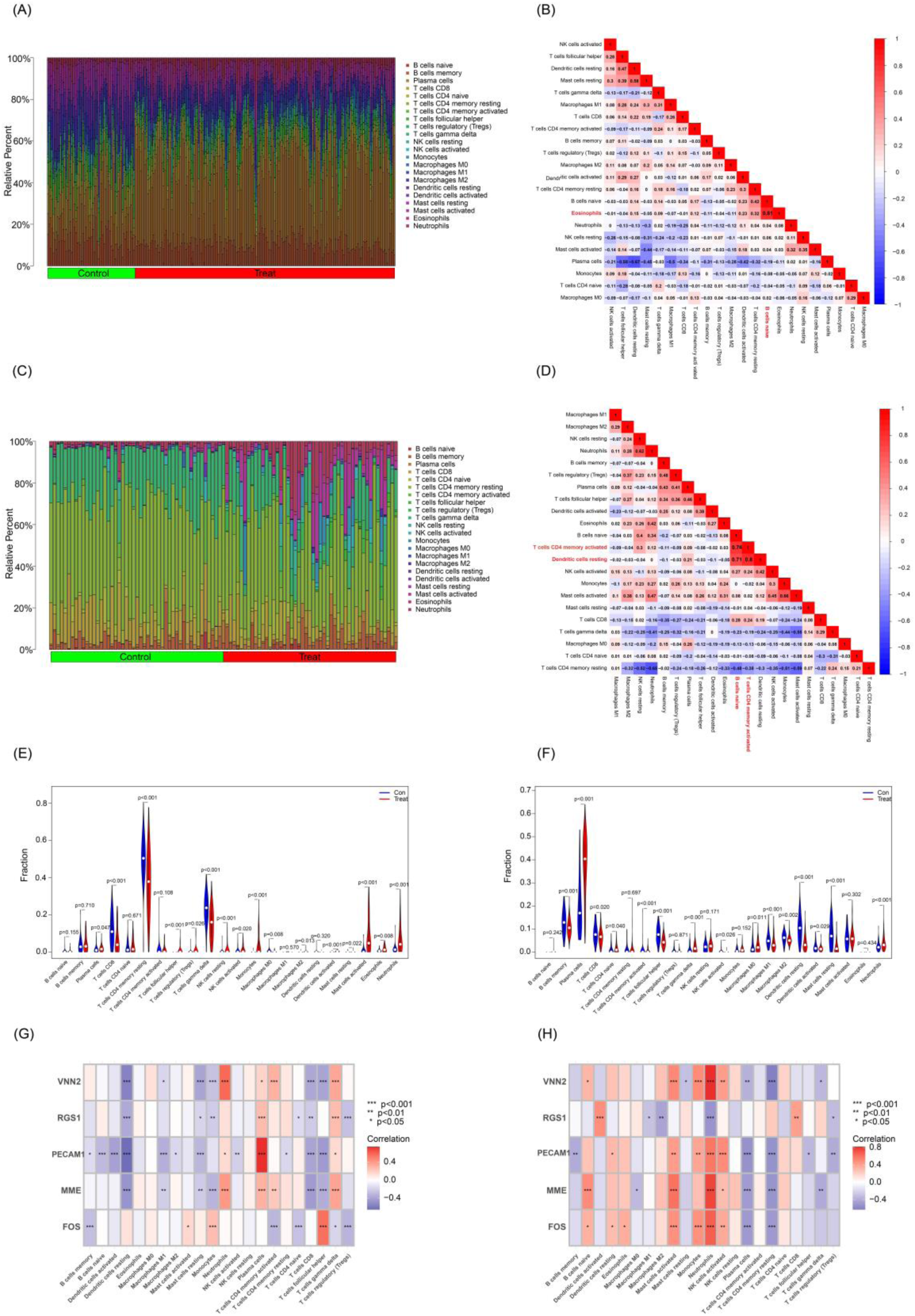
(A, C) Stacked bar chart of the immune cell. The different colors of the rectangular bars in the diagram represent different immune cells, and the length represents the proportion of immune cells. (B, D) The correlation matrix of immune cell proportions. The numbers in the squares represent the correlation coefficients between the corresponding immune cells. (E) Comparison of immune cell scores in patients with periodontitis and a healthy population. (F) Comparison of immune cell fractions in patients with coronary artery disease and a healthy population. The horizontal axis indicates the different immune cells and the vertical axis indicates the proportion of immune cells. T-test was used to compare the disease group with the control group. (G) Correlation between characterized genes and immune cell fractions in patients with periodontitis. (H) Correlation between characteristic genes and immune cell components in patients with coronary artery disease.

In patients with periodontitis and coronary artery disease, there was a significant increase in monocytes, and neutrophils and a significant decrease in activated memory CD4+ T cells (Fig. 8E, F). We next investigated the relationship between the identified signature genes and immune cell components. In periodontitis, *VNN2, PECAM1, MME*, and RGS1 displayed strong negative correlations with CD8+ T cells, while *VNN2* and *MME* showed strong positive correlations with neutrophils (Fig. 8G). Similarly, in CAD, *VNN2, PECAM1*, *MME,* and *FOS* exhibited strong positive correlations with neutrophils, whereas *RGS1* showed a negative correlation (Fig. 8H). These findings suggest that the signature genes may influence immune responses in both periodontitis and CAD.

### 3.7 Clinical validation of RNA-seq analysis

Differential gene expression analyses were performed using “limma” packages. Of the five characterized genes we screened using the machine learning algorithm, four of them were expressed differently in two groups. Specially, the *FOS* gene was confirmed to be highly expressed (logFC > 1.5, P-value = 0.01) in the PC group compared to the C group (Fig. 9). Overall, the expression pattern of the *FOS* gene was highly consistent in RNA sequencing of clinical samples as well as in the analyses of the present study, confirming the potential of the *FOS* gene as a signature gene for coronary heart disease and periodontitis.

**Figure 9.**
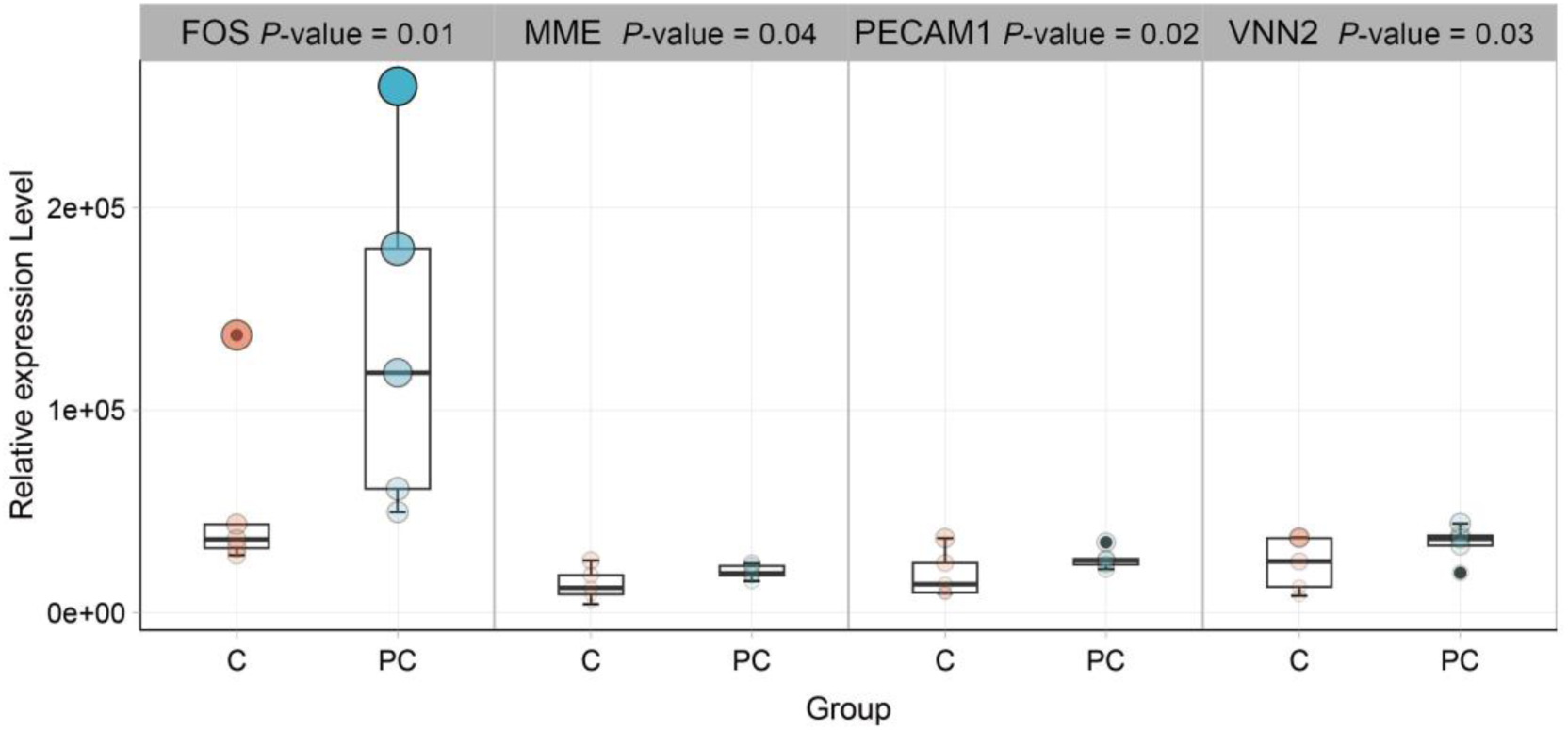
Peripheral blood RNA sequencing analysis of the coronary artery disease group (C group) and the periodontitis combined with coronary artery disease group (PC group). The box plot indicated that *FOS*, *MME*, *PECAM1* and *VNN2* genes were up-regulated in the PC group compared with C group (|logFC| > 1, P-value < 0.05).

## 4 Discussion

Periodontitis and coronary artery disease are both important public health problems, but their correlation remains controversial. In addition to dyslipidemia and metabolic dysfunction, arterial wall inflammation is an important marker of CAD, and the association between CAD and PD may be mediated by PD-induced systemic inflammation because of the high bacterial burden and accumulation of toxic by-products in PD-induced inflammatory connective tissues, and the elevated levels of circulating inflammatory biomarkers are a key response to CAD progression. The importance of inflammation in atherosclerosis is well established and inflammatory markers such as high-sensitivity c-reactive protein (hsCRP) are being used for cardiac risk stratification [26–27]. The inflammatory response can lead to plaque rupture or erosion, setting the stage for a thrombotic response that can lead to myocardial injury or infarction [28]. Alleviating the inflammatory process is an unmet therapeutic need for CAD. With the prevalence of atherosclerosis risk factors such as aging, obesity, and metabolic syndrome, the discovery of new biomarkers and therapeutic targets may be beneficial in the management of this common disease [29–30].

In this study, we integrated the gene expression profiles of periodontitis and coronary artery disease by using bioinformatics, explored the common mechanism between the two from the molecular genetics perspective, explained the potential crosstalk genes, shared pathways, and screened biomarkers with certain significance between periodontitis and coronary artery disease by using the machine learning method, and finally collected clinical samples to perform the RNA-seq test, which verified the screened genes, providing reference for early prevention and diagnosis of patients with the combination of the two diseases.

We extracted genes from modules closely related to periodontitis and coronary artery disease, respectively, and then intersected the module signature genes with DEGs identified by GEO2R to further obtain 48 common DEGs. After GO enrichment analysis, we showed that common DEGs were enriched for biological processes such as response to lipopolysaccharides, response to molecules of bacterial origin, myeloid leukocyte migration, and neutrophil migration; the enriched cellular components were secretory granule membrane, secretory granule lumen, and plasma membrane; and the molecular functions enriched were *CXCR* chemokine receptor binding, pattern recognition receptor activity, and phosphoprotein binding. Here, we further identified five signature genes (*FOS*, *VNN2*, *PECAM1*, *MME*, *RGS1*) associated with inflammation and immune response, and we evaluated the expression of the five signature genes in an external validation set, which showed that the screened signature genes were highly expressed in periodontitis and coronary heart disease, and the AUCs were all greater than 0.7, suggesting that these genes may play a potential role in periodontitis and coronary heart disease. Using RNA-seq experiments on clinical samples, we verified that the expression of *FOS* genes was indeed significantly upregulated in the group with coronary heart disease combined with periodontitis compared to the group with coronary heart disease. *c-Fos* is a transcription factor involved in many signaling pathways, and in view of its prominent role in the severity of atherosclerosis, our study further suggests that *FOS* is likely to act as a biomarker independent of classical such as CRP or white blood cells (WBC), as a core inflammatory marker that influences the onset and progression of coronary heart disease.

The *FOS* gene was originally identified as an oncogene in osteosarcoma, and its importance in inflammation and calcification coincides with known pathological changes in atherosclerosis [31–35]. Neelanjan Ray et al. explored the role of the transcription factor *c-Fos* in lipopolysaccharide (LPS)-induced cytokine responses using mice lacking *c-Fos*. Compared to wild-type controls, *c-Fos* deficient mice showed significantly increased production of tumor necrosis factor (TNF)-a, interleukin *(IL)-6* and *IL-12*, but decreased production of the anti-inflammatory cytokine *IL-10* [36]. (IL)-6 propagates inflammation and stimulates hepatic synthesis of C-reactive proteins and leads to an increase in C-reactive proteins in PD patients [37]. This suggests a novel role for *c-Fos* as an anti-inflammatory transcription factor in oral health and CAD. Meanwhile, N.J. Dun et al. suggested that the differential distribution of *FOS* neurons in hypotensive and hypertensive animals highlights the potential application of *FOS* as a metabolic marker in identifying neuronal networks for specific cardiovascular diseases [38]. The *FOS* gene encodes the transcription factor *FOS* protein, which is involved in the regulation of biological processes such as gene expression and cellular signaling and may be associated with the development of cardiovascular diseases.

The understanding of inflammation in atherosclerosis is currently controversial [33]. This is particularly true for cellular molecules involved in inflammation, as their involvement is usually multifactorial. A second factor is that the application of anti-inflammatory treatments is complex, which is related to the fact that most experimental work has been done using animal models rather than human tissue [34]. For example, in a study originally designed by Aikawa et al. for arthritis treatment, they describe that T5224 (a *Fos/AP-1* inhibitor) significantly suppressed the levels of inflammatory cytokines (*IL-1b*, *IL-6*, and *COMP*) and *MMP3* in vivo and in vitro, suggesting an interesting role in immunosuppression [41]. Similarly, the inhibitory effect of T5224 on *Fos/AP-1* was anti-inflammatory in endotoxin-induced acute kidney injury and in a lipopolysaccharide-induced liver injury model [42]. Zhuang et al. conducted a study on male C57BL/6 mice to investigate immune heterogeneity following myocardial infarction. Mice underwent either myocardial infarction surgery and were monitored for 1 or 7 days, or sham surgery and were monitored for 7 days. Cardiac CD45-positive immune cells were subsequently isolated and subjected to single-cell RNA sequencing analysis. *FOS* protein deficiency attenuates cardiac inflammation by down-regulating *IL-6* and *MCP1* (Monocyte chemotactic protein-1) expression Response. These studies suggest that inhibition of *Fos/AP-1* activity is a potential anti-inflammatory therapeutic approach, and *Fos/AP-1* (activator protein 1) regulation has been identified as a key regulator of pro-inflammatory responses. And enriched *Fos/AP-1* target gene loci were identified in genome-wide association study signals for coronary artery disease and myocardial infarction. Targeting *Fos/AP-1* with the selective inhibitor T5224 attenuated leukocyte infiltration and reduced cardiac dysfunction in a preclinical mouse model of myocardial infarction [43]. In order to identify disease markers and genes associated with atherosclerosis, Patino et al. used the Sequence Analysis of Gene Expression (SAGE) technique to quantify gene expression in circulating monocytes from a limited number of atherosclerotic patients and normal subjects. This comparison showed that, compared with normal controls, the transcriptional levels of a wide range of stress-responsive and inflammatory genes in patient monocytes were higher, especially the *FOS* gene was strongly expressed in patient circulating monocytes. Levels of *FOS* transcripts were increased 8-fold in patients requiring CEA (carotid endarterectomy) compared to controls and were more sensitive and specific for disease severity compared to plasma hsCRP testing [44].

However, the individuals who might benefit from *FOS* treatment have not yet been identified. This calls for the development of a simpler, more sensitive, and more specific *FOS* assay, which would allow for larger prospective clinical trials to determine the clinical utility of *FOS* levels. *FOS* is a responsive transcriptional regulator, a functional property that may make it useful in monitoring disease activity or therapeutic efficacy (45). In addition to this study’s finding that *FOS* expression is elevated in periodontitis conditions, *FOS* may be elevated in other inflammatory conditions, such as rheumatoid arthritis, and needs to be tested in appropriate patient populations [46–47]. *FOS* expression may be equivalent to the coronary calcium score currently used for screening for coronary artery disease [48]. Targeted patient management, based in part on sensitive molecular testing, could provide better insight into atherosclerosis for risk stratification and treatment.

## 5 Conclusion

This study identified five signature genes that were significantly associated with immune cell dysregulation. These genes hold promise for the development of a nomogram-based approach for early diagnosis of both periodontitis and coronary artery disease, potentially informing future research directions for improved diagnosis and treatment strategies in these prevalent conditions. Notably, the prominent upregulation of *FOS* suggests its potential as a key target for future investigations. These insights hold significant implications for improving prevention and diagnostic strategies for individuals affected by both periodontitis and CAD.

## Ethical approval and consent to participate

All the patients’ samples were obtained the informed consent and ethical approval of Renmin Hospital, Hubei University of Medicine.

## Availability of data and materials

Gene expression data from NCBI GEO datasets, GSE10334, GSE66360, GSE6751 and GSE71226 were used in the study. The RNA-Seq data of clinical samples used in this study are available for academic use upon request.

## Authors’ contribution

Shaobin Wang and Wenting Liu supervised the project. Nannan Yang and Kun Wang analyzed the data and drafted the manuscript. Wenqian Yang guided the bioinformatics analysis. Huanhuan Xing helped on bioinformatics analysis. Ying Jin guided the patient samples collection. Hao Zhang, Liguo Tan, Zhen Huang and Qi Chen collected and processed patient’s peripheral blood samples. Wenqian Yang, Wenting Liu and Shaobin Wang revised the manuscript. All authors approved the manuscript.

## References

[1] Kwon T, Lamster IB, Levin L. Current Concepts in the Management of Periodontitis. Int Dent J. 2021;71(6):462–476. doi: 10.1111/idj.12630

[2] Kassebaum NJ, Bernabé E, Dahiya M, Bhandari B, Murray CJ, Marcenes W. Global burden of severe periodontitis in 1990-2010: a systematic review and meta-regression. J Dent Res. 2014;93(11):1045–53. doi: 10.1177/0022034514552491

[3] Peixue Sun, Yuwen Ruan, Chunyan Luo. Research progress on genes related to coronary heart disease.Genomics and Applied Biology J. 2024;43(06):958–966. doi:10.13417/j.gab.043.000958.

[4] Interpretation of the key points of the China Cardiovascular Health and Disease Report 2022. Chinese Cardiovascular Journal. 2023;28 (04): 297–312.

[5] Holmlund A, Holm G, Lind L. Severity of periodontal disease and number of remaining teeth are related to the prevalence of myocardial infarction and hypertension in a study based on 4,254 subjects. J Periodontol. 2006;77(7):1173–8. doi: 10.1902/jop.2006.050233.

[6] Shetty B, Fazal I, Khan SF, Nambiar M, D KI, Prasad R, Raj A. Association between cardiovascular diseases and periodontal disease: more than what meets the eye. Drug Target Insights. 2023;17:31–38. doi: 10.33393/dti.2023.2510.

[7] Sumayin Ngamdu K, Mallawaarachchi I, Dunipace EA, Chuang LH, Jafri SH, Shah NR, Jeong YN, Morrison AR, Bhatt DL. Association Between Periodontal Disease and Cardiovascular Disease (from the NHANES). Am J Cardiol. 2022;178:163–168. doi: 10.1016/j.amjcard.2022.05.028

[8] Sanz M, Marco Del Castillo A, Jepsen S, Gonzalez-Juanatey JR, D’Aiuto F, Bouchard P, Chapple I, Dietrich T, Gotsman I, Graziani F, Herrera D, Loos B, Madianos P, Michel JB, Perel P, Pieske B, Shapira L, Shechter M, Tonetti M, Vlachopoulos C, Wimmer G. Periodontitis and cardiovascular diseases: Consensus report. J Clin Periodontol. 2020;47(3):268–288. doi: 10.1111/jcpe.13189

[9] Joshi C, Bapat R, Anderson W, Dawson D, Hijazi K, Cherukara G. Detection of periodontal microorganisms in coronary atheromatous plaque specimens of myocardial infarction patients: A systematic review and meta-analysis. Trends Cardiovasc Med. 2021;31(1):69–82. doi: 10.1016/j.tcm.2019.12.005

[10] Stryjewska K, Pytko-Polonczyk J, Sagbraaten MS, Sagbraaten SVM, Stryjewski PJ. Prevalence of bacterial and fungal infections the oral cavity in patients with acute myocardial infarction treated with primary coronary intervention. Pol Merkur Lekarski. 2019;47(280):123–127

[11] Edgar R, Domrachev M, Lash AE. Gene Expression Omnibus: NCBI gene expression and hybridization array data repository. Nucleic Acids Res. 2002;30(1):207–10. doi: 10.1093/nar/30.1.207

[12] Demmer RT, Behle JH, Wolf DL, Handfield M, Kebschull M, Celenti R, Pavlidis P, Papapanou PN. Transcriptomes in healthy and diseased gingival tissues. J Periodontol. 2008;79(11):2112–24. doi: 10.1902/jop.2008.080139

[13] Muse ED, Kramer ER, Wang H, Barrett P, Parviz F, Novotny MA, Lasken RS, Jatkoe TA, Oliveira G, Peng H, Lu J, Connelly MC, Schilling K, Rao C, Torkamani A, Topol EJ. A Whole Blood Molecular Signature for Acute Myocardial Infarction. Sci Rep. 2017;7(1):12268. doi: 10.1038/s41598-017-12166-0

[14] Langfelder P, Horvath S. WGCNA: an R package for weighted correlation network analysis. BMC Bioinformatics. 2008;9:559. doi: 10.1186/1471-2105-9-559

[15] Barrett T, Wilhite SE, Ledoux P, Evangelista C, Kim IF, Tomashevsky M, Marshall KA, Phillippy KH, Sherman PM, Holko M, Yefanov A, Lee H, Zhang N, Robertson CL, Serova N, Davis S, Soboleva A. NCBI GEO: archive for functional genomics data sets--update. Nucleic Acids Res. 2013;41(Database issue):D991–5. doi: 10.1093/nar/gks1193.

[16] Bu D, Luo H, Huo P, Wang Z, Zhang S, He Z, Wu Y, Zhao L, Liu J, Guo J, Fang S, Cao W, Yi L, Zhao Y, Kong L. KOBAS-i: intelligent prioritization and exploratory visualization of biological functions for gene enrichment analysis. Nucleic Acids Res. 2021;49(W1):W317–W325. doi: 10.1093/nar/gkab447

[17] Breiman L. Random Forests. Machine Learning, 2001,45(1):5–32

[18] Jeong B, Cho H, Kim J, Kwon SK, Hong S, Lee C, Kim T, Park MS, Hong S, Heo TY. Comparison between Statistical Models and Machine Learning Methods on Classification for Highly Imbalanced Multiclass Kidney Data. Diagnostics(Basel). 2020;10(6):415. doi: 10.3390/diagnostics10060415

[19] Huang S, Cai N, Pacheco PP, Narrandes S, Wang Y, Xu W. Applications of Support Vector Machine (SVM) Learning in Cancer Genomics. Cancer Genomics Proteomics. 2018;15(1):41–51. doi: 10.21873/cgp.20063

[20] Dey A, Hankey Giblin PA. Insights into Macrophage Heterogeneity and Cytokine-Induced Neuroinflammation in Major Depressive Disorder. Pharmaceuticals (Basel). 2018;11(3):64. doi: 10.3390/ph11030064

[21] Lee CS, Conway C. The role of generalized linear models in handling cost and count data. Eur J Cardiovasc Nurs. 2022;21(4):392–398. doi:10.1093/eurjcn/zvac002

[22] Pan X, Jin X, Wang J, Hu Q, Dai B. Placenta inflammation is closely associated with gestational diabetes mellitus. Am J Transl Res. 2021;13(5):4068–4079

[23] Newman AM, Liu CL, Green MR, Gentles AJ, Feng W, Xu Y, Hoang CD, Diehn M, Alizadeh AA. Robust enumeration of cell subsets from tissue expression profiles. Nat Methods. 2015;12(5):453–7. doi: 10.1038/nmeth.3337

[24] Hu K. Become Competent within One Day in Generating Boxplots and Violin Plots for a Novice without Prior R Experience. Methods Protoc. 2020;3(4):64. doi: 10.3390/mps3040064

[25] Anders S, Pyl PT, Huber W. HTSeq--a Python framework to work with high-throughput sequencing data. Bioinformatics. 2015 Jan 15;31(2):166–9. doi:10.1093/bioinformatics/btu638

[26] Ross R. Atherosclerosis--an inflammatory disease. N Engl J Med. 1999;340(2):115–26. doi: 10.1056/NEJM199901143400207

[27] Libby P, Ridker PM, Maseri A. Inflammation and atherosclerosis. Circulation. 2002; 105(9):1135–43. doi: 10.1161/hc0902.104353

[28] Harrington RA. Targeting Inflammation in Coronary Artery Disease. N Engl J Med. 2017;377(12):1197–1198. doi: 10.1056/NEJMe1709904

[29] Chaves PH, Kuller LH, O’Leary DH, Manolio TA, Newman AB; Cardiovascular Health Study. Subclinical cardiovascular disease in older adults: insights from the Cardiovascular Health Study. Am J Geriatr Cardiol. 2004;13(3):137–51. doi: 10.1111/j.1076-7460.2004.02120

[30] Willerson JT, Ridker PM. Inflammation as a cardiovascular risk factor. Circulation. 2004;109(21 Suppl 1):II2–10. doi:10.1161/01.CIR.0000129535.04194.38

[31] Ransone LJ, Verma IM. Nuclear proto-oncogenes fos and jun. Annu Rev Cell Biol. 1990;6:539–57. doi: 10.1146/annurev.cb.06.110190.002543

[32] Johnson RS, Spiegelman BM, Papaioannou V. Pleiotropic effects of a null mutation in the c-fos proto-oncogene. Cell. 1992;71(4):577–86. doi: 10.1016/0092-8674(92)90592-z

[33] Wang ZQ, Ovitt C, Grigoriadis AE, Möhle-Steinlein U, Rüther U, Wagner EF. Bone and haematopoietic defects in mice lacking c-fos. Nature. 1992;360(6406):741–5. doi: 10.1038/360741a0

[34] Liebermann DA, Gregory B, Hoffman B. AP-1 (Fos/Jun) transcription factors in hematopoietic differentiation and apoptosis. Int J Oncol. 1998;12(3):685–700. doi: 10.3892/ijo.12.3.685

[35] Doherty TM, Fitzpatrick LA, Shaheen A, Rajavashisth TB, Detrano RC. Genetic determinants of arterial calcification associated with atherosclerosis. Mayo Clin Proc. 2004;79(2):197–210. doi: 10.4065/79.2.197

[36] Ray N, Kuwahara M, Takada Y, Maruyama K, Kawaguchi T, Tsubone H, Ishikawa H, Matsuo K. c-Fos suppresses systemic inflammatory response to endotoxin. Int Immunol. 2006;18(5):671–7. doi: 10.1093/intimm/dxl004

[37] Sancé au J, Kaisho T, Hirano T, Wietzerbin J. Triggering of the human interleukin-6 gene by interferon-gamma and tumor necrosis factor-alpha in monocytic cells involves cooperation between interferon regulatory factor-1, NF kappa B, and Sp1 transcription factors. J Biol Chem. 1995;270(46):27920–31. doi: 10.1074/jbc.270.46.27920

[38] Dun NJ, Dun SL, Shen E, Tang H, Huang R, Chiu TH. c-fos expression as a marker of central cardiovascular neurons. Biol Signals. 1995;4(3):117–23. doi: 10.1159/000109431

[39] Libby P, Hansson GK. From Focal Lipid Storage to Systemic Inflammation: JACC Review Topic of the Week. J Am Coll Cardiol. 2019;74(12):1594–1607. doi:10.1016/j.jacc.2019.07.061

[40] Bhaskar V, Yin J, Mirza AM, Phan D, Vanegas S, Issafras H, Michelson K, Hunter JJ, Kantak SS. Monoclonal antibodies targeting IL-1 beta reduce biomarkers of atherosclerosis in vitro and inhibit atherosclerotic plaque formation in Apolipoprotein E-deficient mice. Atherosclerosis. 2011;216(2):313–20. doi: 10.1016/j.atherosclerosis

[41] Aikawa Y, Morimoto K, Yamamoto T, Chaki H, Hashiramoto A, Narita H, Hirono S, Shiozawa S. Treatment of arthritis with a selective inhibitor of c-Fos/activator protein-1. Nat Biotechnol. 2008;26:817–823. doi: 10.1038/nbt1412

[42] Miyazaki H, Morishita J, Ueki M, Nishina K, Shiozawa S, Maekawa N. The effects of a selective inhibitor of c-Fos/activator protein-1 on endotoxin-induced acute kidney injury in mice. BMC Nephrol. 2012;13:153. doi: 10.1186/1471-2369-13-153

[43] Zhuang L, Wang Y, Chen Z, Li Z, Wang Z, Jia K, Zhao J, Zhang H, Xie H, Lu L, Chen K, Chen L, Fukuda K, Sano M, Zhang R, Liu J, Yan X. Global Characteristics and Dynamics of Single Immune Cells After Myocardial Infarction. J Am Heart Assoc. 2022;11(24):e027228. doi: 10.1161/JAHA.122.027228

[44] Patino WD, Mian OY, Kang JG, Matoba S, Bartlett LD, Holbrook B, Trout HH 3rd, Kozloff L, Hwang PM. Circulating transcriptome reveals markers of atherosclerosis. Proc Natl Acad Sci USA. 2005;102(9):3423–8. doi: 10.1073/pnas.0408032102

[45] Ransone LJ, Verma IM. Nuclear proto-oncogenes fos and jun. Annu Rev Cell Biol. 1990;6:539–57. doi: 10.1146/annurev.cb.06.110190.002543

[46] Ridker PM. Clinical application of C-reactive protein for cardiovascular disease detection and prevention. Circulation. 2003;107(3):363–9. doi:10.1164/01.cir.0000053730.47739.3c

[47] Onda K, Rimbara E, Hirano T, Oka K, Abe H, Tahara K, Takanashi H, Tsuboi N, Niitsuma T, Hayashi T. Role of mRNA expression of transcription factors in glucocorticoid sensitivity of peripheral blood mononuclear cells and disease state in rheumatoid arthritis. J Rheumatol. 2004;31(3):464–9

[48] Greenland P, LaBree L, Azen SP, Doherty TM, Detrano RC. Coronary artery calcium score combined with Framingham score for risk prediction in asymptomatic individuals. JAMA. 2004;291(2):210–5. doi: 10.1001/jama.291.2.210

